# The EHR Density Index: A new method to control for EHR data inconsistency across patients

**DOI:** 10.64898/2026.08.03.26359595

**Authors:** Abhishek Bhatia, Sydney Lash, Tomas McIntee, Emily R. Pfaff

## Abstract

Electronic health record (EHR) data vary substantially in documentation density across patients, independent of disease burden. Existing tools such as the Charlson Comorbidity Index (CCI) and Elixhauser Comorbidity Index measure disease burden but do not capture differences in data volume, leaving a common source of bias unaddressed in EHR-based analyses. To address this gap, we developed the EHR Density Index (EDI), which characterizes the quantity, depth, and breadth of EHR data per patient per year, normalized by utilization patterns, using records from 24,987 adult patients at UNC Health (2018–2024). The EDI combines a utilization cluster assigned via Gaussian Mixture Model with within-cluster residuals quantifying documentation volume across four clinical domains. Four interpretable clusters emerged; while CCI predicted cluster membership, its associations with within-cluster residuals were weak, confirming the EDI captures dimensions of the patient record distinct from disease burden. The EDI is intended as a covariate to address documentation density as a source of confounding in real-world data-driven research.

## Introduction

Effective use of electronic health record (EHR) data for research requires understanding its limitations and appropriate uses. EHR data falls into the category of “real-world data” (RWD), or data that are not collected expressly for the purpose of research. As RWD, EHR data may have greater levels of missingness and inconsistency than research data. While those challenges do not diminish EHR data’s utility, they do require careful consideration to ensure that research results derived from EHR data are valid, robust, reproducible, and generalizable.

Every research study using EHR data faces the challenge of appropriately handling this characteristic data unevenness when designing analyses. Patients can have widely varying amounts of information available in their EHRs; for instance, it is well established that sicker patients tend to have greater quantities of data than less sick patients.^1–3^ The Charlson Comorbidity Index (CCI)^4,5^ and the Elixhauser Comorbidity Index^6^ are frequently used as covariates in EHR data analyses to attempt to control for this bias, serving as proxies for disease burden. However, it is not always possible or appropriate to calculate the CCI or Elixhauser for each patient in a cohort; calculating a comorbidity index for patients with few encounters, sparse data, short histories with a health system, or who only see certain specialists can be misleading. Moreover, as the CCI was designed to measure mortality risk and not as a multimorbidity measure, it emphasizes conditions associated with poorer survival, and thus may not be well-suited to all analyses.^7,8^ Finally, comorbidity indexes can measure whether a patient is sick, but lack information about how and how often patients interact with the health care system over time, or whether their records are unusually deep or sparse. There is thus a need for a new index to serve a similar analytical purpose to the CCI and Elixhauser, but tuned specifically to volumetric idiosyncrasies of EHR data.

We posit that an index measuring *EHR density*, or the amount of data packed into a patient’s healthcare encounters, bound by calendar years and normalized by utilization patterns, may address some of the analytical downsides of using the CCI and Elixhauser indexes alone. Our intention is not to displace the CCI or Elixhauser indexes, which remain well-vetted and useful measures, but rather to give researchers an additional option to control for uneven data in cohort definitions or analysis plans. Here we describe our effort to create the EHR Density Index (EDI), intended for use as a covariate in downstream analyses alongside measures of comorbidity.

## Methods

### Study population

We received full medical records for 25,000 randomly selected patients from UNC Health, provisioned in the OMOP common data model. Selected patients were ≥18 years of age as of the query date and had at least two encounters with UNC Health between 1/1/2018 and 12/1/2024. 13 patients were eliminated on the basis of outlier age (≥100) or visit dates in the extreme past (likely a data quality issue), leaving a sample of 24,987 for analysis.

### Unified person-date table

To consolidate clinical activity across all OMOP domains, we built a unified person-date table from seven source tables: visit_occurrence, condition_occurrence, drug_exposure, measurement, procedure_occurrence, and person. For each domain, we extracted the relevant date column (e.g., condition_start_date, drug_exposure_start_date) and standardized it to a common date field, then aggregated concept identifiers into arrays per person-date to preserve all clinical information while maintaining a one-row-per-person-date grain. We exploded multi-day visits into daily records so that each hospitalization day was counted, and we flagged inpatient days where the OMOP visit concept ID was either Inpatient Visit (9201) or Emergency Room and Inpatient Visit (262). The resulting person-date spine included all dates with any clinical activity — both formal visit days and days with documented clinical concepts but no recorded visit (e.g., standalone laboratory orders or medication refills).

Our ultimate goal was to create a data density metric that consisted of two components: (1) a utilization cluster, to allow us to normalize each patient’s data density per their overall rate of healthcare utilization and (2) a “density vector,” which serves to quantify the data available for each patient across four clinical domains for each calendar year: conditions, drugs, measurements, and procedures.

### Utilization-based clustering

#### Feature construction

We aggregated the daily table to person-calendar year level, yielding features that captured three core dimensions of utilization: volume (total visit count per year), intensity (inpatient visit count and total hospitalized days per year), and temporal regularity of care (how evenly spaced a patient’s visits were throughout the year).

We quantified visit regularity using a continuous irregularity score. For each person-year with at least two visits, we calculated inter-visit gaps and measured how much those gaps deviated from the patient’s own average gap. We computed both an L1 (mean absolute deviation) and L2 (root mean squared deviation) version of this score; both are zero for perfectly evenly spaced visits and increase with greater irregularity. We also recorded whether each person-year had enough visits (≥3) to meaningfully assess regularity; patients with fewer than three visits received null irregularity scores.

#### Gaussian mixture model

We identified utilization clusters using a Gaussian Mixture Model (GMM) with four components, fit to the person-year utilization features (visit count, inpatient visit count, hospitalized days, irregularity L1 and L2 scores, and a binary indicator for whether regularity could be assessed). GMMs accommodate non-spherical cluster shapes and provide probabilistic assignments, allowing us to quantify uncertainty for each patient’s classification. We created a stratified training sample (one randomly selected year per person) to prevent patients with longer records from dominating model fitting, then applied a standard preprocessing pipeline — winsorizing extreme outliers at the 99th percentile, log-transforming skewed count features, and standardizing all features to zero mean and unit variance (full preprocessing details in Supplementary Methods). We fit the model with tied covariance (a shared covariance matrix across components) and 10 random initializations, selecting the solution with the highest log-likelihood.

We evaluated three alternative clustering approaches as sensitivity analyses: two rule-based classifiers (one with fixed thresholds and one with adaptively tuned thresholds, each yielding seven clusters) and a seven-component GMM. Details are provided in the Supplementary Methods.

#### Cluster interpretation

The four resulting clusters corresponded to interpretable utilization clusters, which our study team named after manually examining the patient-year records included in each. The clusters are: High Inpatient (patients with multiple or extended hospitalizations), Moderate Inpatient (patients with occasional inpatient stays), Outpatient Regular (patients seen in ambulatory settings at regularly-spaced encounters), and Outpatient Irregular (patients with minimal, irregularly-spaced outpatient encounters). For each patient-year, the model produced a posterior probability across all four clusters; we used the maximum posterior probability as a confidence score and the Shannon entropy of the distribution as an uncertainty measure.

### Data Density Vectors

#### Domain-level concept counts

To characterize the amount and breadth of clinical documentation available for each patient-year, we computed data volume metrics from the four non-visit OMOP domains: conditions, drugs, procedures, and measurements. These domain counts were calculated independently of the utilization features used for clustering.

For each person-year and domain, we calculated the total number of concept occurrences (non-unique count, reflecting volume of documentation regardless of number of unique concepts). We also computed the total data points across all four domains for each patient-year. These metrics capture the depth of a patient’s clinical record and vary substantially across utilization clusters.

#### Within-cluster domain residuals

To assess how a patient’s documentation volume deviates from others with similar utilization patterns, we computed within-cluster residuals for each of the four unique domain counts for each patient-year. For each domain, we mean-centered the count within each GMM-4 cluster, yielding a signed residual: positive values indicate a patient has more concept instances in that domain than the cluster average, and negative values indicate fewer. We also computed unsigned (absolute) residuals to capture the magnitude of deviation regardless of direction. These residuals are in raw count space (not standardized), and because the domain counts were not used in the clustering solution, the residuals reflect documentation density variation that is independent of cluster assignment. We termed the resulting array of four residuals for each patient-year our “density vector.”

#### Association between disease burden and data volume

We ran additional analyses to determine the degree to which our density vectors are independent of patient disease burden, defined as the cumulative maximum CCI for each patient. We computed pooled Spearman rank correlations between total CCI and each domain residual (both signed and unsigned) across all clusters and person-years, providing a global measure of the association between disease severity and documentation deviation from cluster norms. We stratified the same correlations within each utilization cluster to check for Simpson’s paradox–that is, whether the pooled association obscures opposing or heterogeneous effects across clusters (Bonferroni-corrected alpha = 0.05 / 4 = 0.0125). We fit OLS interaction models of the form *residual ∼ CCI × cluster* with clustered standard errors by patient to test whether the CCI-residual slope differs across clusters; for unsigned outcomes with high skewness (>1.0), we applied a log(1 + x) transformation.

Finally, we fit a multinomial logistic regression predicting cluster assignment from CCI, with Outpatient Regular as the reference category, to estimate the odds of belonging to each higher-utilization cluster per unit increase in CCI.

Because patients contribute multiple person-years, standard p-values are inflated by non-independence; we used clustered standard errors in OLS models and emphasize effect sizes (Spearman ρ, odds ratios) over significance thresholds throughout.

## Results

### Patient Characteristics

The cohort consists of 24,987 patients. Here, we present breakdowns for patients stratified by utilization cluster (Table 1a) and cross-domain residuals (Table 1b)–i.e., the average residual across all four clinical domains.

**Table 1a.** Patient characteristics by utilization cluster, 2022.

| Variable | Utilization Cluster |  |  |  |  |
| --- | --- | --- | --- | --- | --- |
|  | All patients<br>(n=14826) | High inpatient (n=406) | Moderate inpatient<br>(n= 726) | Outpatient regular<br>(n= 9199) | Outpatient irregular<br>(n=4495) |
| Age (as of 1/1/2022) |  |  |  |  |  |
| 18-25 | 1702 (11.5) | 18 (4.4) | 58 (8) | 834 (9.1) | 792 (17.6) |
| 26-35 | 1910 (12.9) | 19 (4.7) | 169 (23.3) | 971 (10.6) | 751 (16.7) |
| 36-45 | 1931 (13) | 19 (4.7) | 102 (14) | 1112 (12.1) | 698 (15.5) |
| 46-55 | 2314 (15.6) | 46 (11.3) | 66 (9.1) | 1509 (16.4) | 693 (15.4) |
| 56-65 | 2644 (17.8) | 68 (16.7) | 76 (10.5) | 1803 (19.6) | 697 (15.5) |
| Over 65 | 4325 (29.2) | 236 (58.1) | 255 (35.1) | 2970 (32.3) | 864 (19.2) |
| Sex |  |  |  |  |  |
| Female | 9028 (60.9) | 211 (52) | 490 (67.5) | 5720 (62.2) | 2607 (58) |
| Male | 5798 (39.1) | 195 (48) | 236 (32.5) | 3479 (37.8) | 1888 (42) |
| Race |  |  |  |  |  |
| Black or African-American | 3081 (20.8) | 111 (27.3) | 182 (25.1) | 1856 (20.2) | 932 (20.7) |
| Other or Unknown | 1948 (13.1) | 29 (7.1) | 89 (12.3) | 1113 (12.1) | 717 (16) |
| White | 9797 (66.1) | 266 (65.5) | 455 (62.7) | 6230 (67.7) | 2846 (63.3) |
| Ethnicity |  |  |  |  |  |
| Hispanic or Latino | 987 (6.7) | 14 (3.4) | 68 (9.4) | 555 (6) | 350 (7.8) |
| Not Hispanic or Latino | 13839 (93.3) | 392 (96.6) | 658 (90.6) | 8644 (94) | 4145 (92.2) |
| Maximum CCI (as of 1/1/2022) |  |  |  |  |  |
| 0 | 10341 (69.7) | 78 (19.2) | 391 (53.9) | 6001 (65.2) | 3871 (86.1) |
| 1 | 1935 (13.1) | 64 (15.8) | 98 (13.5) | 1427 (15.5) | 346 (7.7) |
| 2 | 1175 (7.9) | 75 (18.5) | 76 (10.5) | 869 (9.4) | 155 (3.4) |
| 3 | 566 (3.8) | 74 (18.2) | 64 (8.8) | 365 (4) | 63 (1.4) |
| 4 | 348 (2.3) | 52 (12.8) | 39 (5.4) | 229 (2.5) | 28 (0.6) |
| 5 | 196 (1.3) | 20 (4.9) | 22 (3) | 141 (1.5) | 13 (0.3) |
| 6+ | 265 (1.8) | 43 (10.6) | 36 (5) | 167 (1.8) | 19 (0.4) |
| Years with UNC data (between 2018-2024) |  |  |  |  |  |
| 1 | 245 (1.7) | 16 (3.9) | 40 (5.5) | 159 (1.7) | 30 (0.7) |
| 2 | 1412 (9.5) | 35 (8.6) | 79 (10.9) | 532 (5.8) | 766 (17) |
| 3 | 2593 (17.5) | 60 (14.8) | 102 (14) | 1288 (14) | 1143 (25.4) |
| 4 | 2285 (15.4) | 54 (13.3) | 126 (17.4) | 1191 (12.9) | 914 (20.3) |
| 5 | 2147 (14.5) | 71 (17.5) | 103 (14.2) | 1233 (13.4) | 740 (16.5) |
| 6 | 2083 (14) | 73 (18) | 100 (13.8) | 1416 (15.4) | 494 (11) |
| 7 | 4061 (27.4) | 97 (23.9) | 176 (24.2) | 3380 (36.7) | 408 (9.1) |

**Table 1b.** Patient characteristics by average cross-domain residual, 2022. For the purposes of succinctly summarizing patient characteristics, we averaged the residuals for each of the four clinical domains represented in the patient-year density vectors (conditions, drugs, measurements, and procedures) to determine whether a patient’s 2022 *overall* residual is above or below the population average within that patient’s 2022 utilization cluster.

| Variable | Residuals |  |  |
| --- | --- | --- | --- |
|  | All patients (n=14826) | Residuals above average for cluster (n=5125) | Residuals below average for cluster (n=9701) |
| Age (as of 1/1/2022) |  |  |  |
| 18-25 | 1702 (11.5) | 531 (10.4) | 1171 (12.1) |
| 26-35 | 1910 (12.9) | 594 (11.6) | 1316 (13.6) |
| 36-45 | 1931 (13) | 636 (12.4) | 1295 (13.3) |
| 46-55 | 2314 (15.6) | 746 (14.6) | 1568 (16.2) |
| 56-65 | 2644 (17.8) | 939 (18.3) | 1705 (17.6) |
| Over 65 | 4325 (29.2) | 1679 (32.8) | 2646 (27.3) |
| Sex |  |  |  |
| Female | 9028 (60.9) | 3028 (59.1) | 6000 (61.8) |
| Male | 5798 (39.1) | 2097 (40.9) | 3701 (38.2) |
| Race |  |  |  |
| Black or African-American | 3081 (20.8) | 1226 (23.9) | 1855 (19.1) |
| Other or Unknown | 1948 (13.1) | 541 (10.6) | 1407 (14.5) |
| White | 9797 (66.1) | 3358 (65.5) | 6439 (66.4) |
| Ethnicity |  |  |  |
| Hispanic or Latino | 987 (6.7) | 304 (5.9) | 683 (7) |
| Not Hispanic or Latino | 13839 (93.3) | 4821 (94.1) | 9018 (93) |
| Maximum CCI (as of 1/1/2022) |  |  |  |
| 0 | 10341 (69.7) | 3059 (59.7) | 7282 (75.1) |
| 1 | 1935 (13.1) | 762 (14.9) | 1173 (12.1) |
| 2 | 1175 (7.9) | 556 (10.8) | 619 (6.4) |
| 3 | 566 (3.8) | 284 (5.5) | 282 (2.9) |
| 4 | 348 (2.3) | 168 (3.3) | 180 (1.9) |
| 5 | 196 (1.3) | 119 (2.3) | 77 (0.8) |
| 6+ | 265 (1.8) | 177 (3.5) | 88 (0.9) |
| Years with UNC data (between 2018-2024) |  |  |  |
| 1 | 245 (1.7) | 79 (1.5) | 166 (1.7) |
| 2 | 1412 (9.5) | 459 (9) | 953 (9.8) |
| 3 | 2593 (17.5) | 766 (14.9) | 1827 (18.8) |
| 4 | 2285 (15.4) | 714 (13.9) | 1571 (16.2) |
| 5 | 2147 (14.5) | 699 (13.6) | 1448 (14.9) |
| 6 | 2083 (14) | 695 (13.6) | 1388 (14.3) |
| 7 | 4061 (27.4) | 1713 (33.4) | 2348 (24.2) |

Because utilization clusters and residuals are calculated at the patient-year level, we have arbitrarily chosen to present data from 2022. As such, the tables below are limited to patients in our cohort with 2022 data (*n =* 14,826).

### The EHR Density Index (EDI)

EDIs are calculated on a per-patient, per-year basis. The EDI is not a single number, but rather a vector of values that can be used in different ways. A fictitious example of three EDIs for two unique patients is shown in Table 2, below.

**Table 2.** Sample EDIs.

| Record identifiers |  | EDI values |  |  |  |  |  |
| --- | --- | --- | --- | --- | --- | --- | --- |
| Patient ID | Year | Cluster | Condition Residual | Medication residual | Measurement residual | Procedure residual | Mean residual |
| 111 | 2022 | Outpatient irregular | -0.08 | -0.60 | 0.01 | -0.03 | -0.18 |
| 111 | 2023 | Outpatient regular | 0.02 | -0.3 | 0.03 | -0.03 | -0.07 |
| 222 | 2025 | High inpatient | 1.21 | 0.02 | 0.93 | 0.87 | 0.76 |

It is intended that any analysis using the EDI would use both the cluster and residual components. However, a user could choose to simplify by using the mean residual rather than accounting for each domain separately. Alternatively, a study focused exclusively on (e.g.) medications may choose to only use the medication residual.

### Relating visit frequency, utilization, and density

### Within-Cluster Domain Residuals

### Disease Burden Association Analyses

As expected, CCI is associated with utilization cluster assignment. While not universally true, patients with high utilization (e.g., the High Inpatient cluster) are more likely to have a higher disease burden.

CCI is also associated with several domain residuals, but weakly. When we look at the association between CCI and domain residuals regardless of cluster, the value of Spearman’s ρ is negligible or weakly positive. (Full details available in Supplemental Results.) When stratified by cluster, we see evidence of Simpson’s paradox, in which relationships obscured by the pooled analysis are evident in the stratified analysis. However, even these relationships (detailed in Table 3, below) are relatively weak, indicating that CCI only modestly tracks within-cluster density deviation. Patients in the same cluster, and even with similar CCI can still have very different data density.

**Table 3.** Stratified Spearman’s ρ shows mostly significant, but weak relationships between CCI and domain residuals.

| Residual type | Domain | Cluster | Patient Years | Unique Patients | Spearman's $\rho$ | p_value |
| --- | --- | --- | --- | --- | --- | --- |
| Signed | Condition | Moderate Inpatient | 4800 | 3968 | 0.3165 | 0.0000 |
| Signed | Condition | Outpatient Irregular | 41978 | 18582 | 0.0304 | 0.0000 |
| Signed | Condition | Outpatient Regular (ref) | 59839 | 20874 | 0.1637 | 0.0000 |
| Signed | Condition | High Inpatient | 2535 | 1968 | 0.3247 | 0.0000 |
| Signed | Medication | Moderate Inpatient | 4800 | 3968 | 0.2181 | 0.0000 |
| Signed | Medication | Outpatient Irregular | 41978 | 18582 | -0.0108 | 0.0263 |
| Signed | Medication | Outpatient Regular (ref) | 59839 | 20874 | 0.1269 | 0.0000 |
| Signed | Medication | High Inpatient | 2535 | 1968 | 0.1796 | 0.0000 |
| Signed | Procedure | Moderate Inpatient | 4800 | 3968 | 0.132 | 0.0000 |
| Signed | Procedure | Outpatient Irregular | 41978 | 18582 | 0.017 | 0.0005 |
| Signed | Procedure | Outpatient Regular (ref) | 59839 | 20874 | 0.1329 | 0.0000 |
| Signed | Procedure | High Inpatient | 2535 | 1968 | 0.1611 | 0.0000 |
| Signed | Measurement | Moderate Inpatient | 4800 | 3968 | 0.2691 | 0.0000 |
| Signed | Measurement | Outpatient Irregular | 41978 | 18582 | 0.0069 | 0.1590 |
| Signed | Measurement | Outpatient Regular (ref) | 59839 | 20874 | 0.1325 | 0.0000 |
| Signed | Measurement | High Inpatient | 2535 | 1968 | 0.3042 | 0.0000 |
| Unsigned | Condition | Moderate Inpatient | 4800 | 3968 | -0.0477 | 0.0009 |
| Unsigned | Condition | Outpatient Irregular | 41978 | 18582 | 0.0448 | 0.0000 |
| Unsigned | Condition | Outpatient Regular (ref) | 59839 | 20874 | 0.0787 | 0.0000 |
| Unsigned | Condition | High Inpatient | 2535 | 1968 | -0.142 | 0.0000 |
| Unsigned | Medication | Moderate Inpatient | 4800 | 3968 | 0.0124 | 0.3901 |
| Unsigned | Medication | Outpatient Irregular | 41978 | 18582 | 0.0445 | 0.0000 |
| Unsigned | Medication | Outpatient Regular (ref) | 59839 | 20874 | 0.0626 | 0.0000 |
| Unsigned | Medication | High Inpatient | 2535 | 1968 | -0.0668 | 0.0008 |
| Unsigned | Procedure | Moderate Inpatient | 4800 | 3968 | -0.0136 | 0.3457 |
| Unsigned | Procedure | Outpatient Irregular | 41978 | 18582 | 0.022 | 0.0000 |
| Unsigned | Procedure | Outpatient Regular (ref) | 59839 | 20874 | 0.0833 | 0.0000 |
| Unsigned | Procedure | High Inpatient | 2535 | 1968 | 0.0213 | 0.2837 |
| Unsigned | Measurement | Moderate Inpatient | 4800 | 3968 | 0.031 | 0.0319 |
| Unsigned | Measurement | Outpatient Irregular | 41978 | 18582 | 0.0271 | 0.0000 |
| Unsigned | Measurement | Outpatient Regular (ref) | 59839 | 20874 | 0.1048 | 0.0000 |
| Unsigned | Measurement | High Inpatient | 2535 | 1968 | -0.104 | 0.0000 |

## Discussion

In this study, we set out to develop an index to be used as a covariate to control for bias resulting from variable EHR data quantity, accounting for utilization, depth, breadth, and longitudinality. To meet this need, we developed the EHR Density Index, or EDI, which is intended for use alongside time-tested measures of disease burden such as the CCI and the Elixhauser Indexes.

The EDI accounts for *utilization* through assignment of a utilization cluster to each patient-year record, reflecting the pattern of that patient-year’s interactions with the health care system. It accounts for *depth* by calculating residuals, or the degree to which the quantity of data in a given patient-year record deviates from the mean in the same utilization cluster. It accounts for *breadth* by calculating separate residuals across four data domains: conditions, medications, measurements, and procedures. It accounts for longitudinality by calculating a separate EDI for each patient for each calendar year, thus acknowledging that patients, health utilization, and overall health change over time.

We found relationships between components of the EDI and the CCI, suggesting an expected association of disease burden and utilization cluster. The associations between CCI and our domain residuals were present, but weak. As shown in Figure 2, density is heterogenous within clusters, making the story more complex than “sicker patient = denser data.” To illustrate why the EDI measures something quite different than the CCI or Elixhauser Index: Imagine a patient with a high disease burden, but who only visits UNC Health for annual dermatology visits. A CCI or Elixhauser Index calculated for this patient would be likely to be artificially low due to missing data. In the EDI, they would likely be assigned to the Outpatient Irregular cluster, and their residuals would only be valid in the context of their dermatological concerns. Despite not having the whole picture of this patient’s health, their EDI is an accurate reflection of their relationship with UNC Health. It is not, however, a proxy for their disease burden. For this reason, we do not propose the EHR Density Index as a replacement for the CCI or Elixhauser Index. Rather, we see it as an accompaniment.

**Figure 1.**
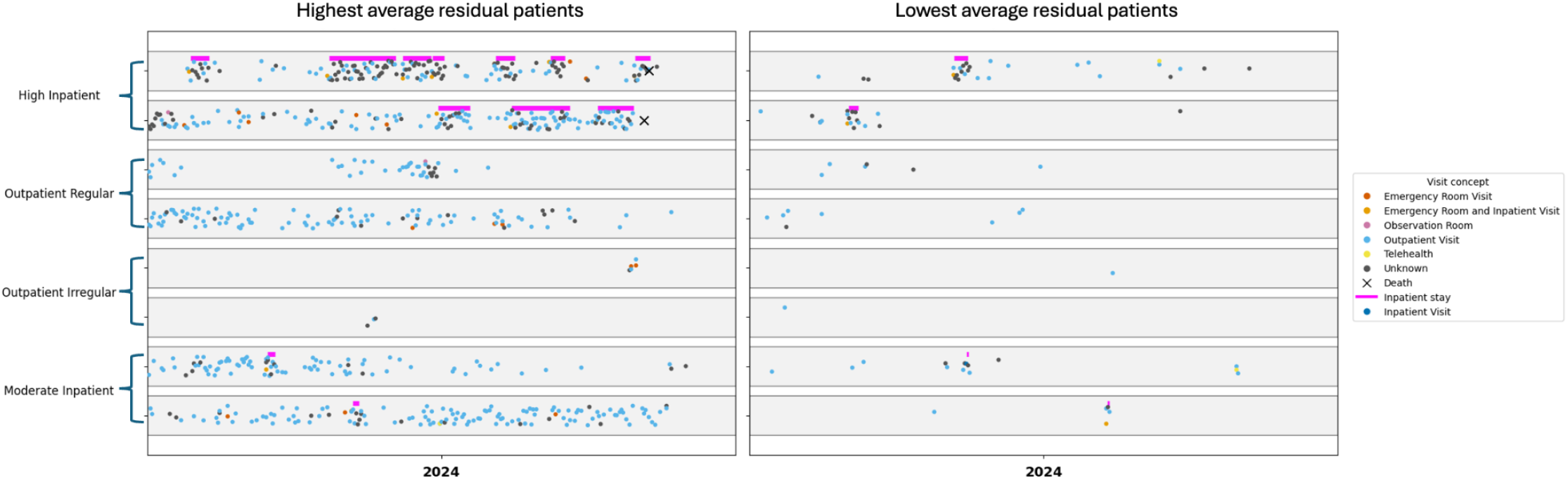
Visualizing visit frequency for high and low density patients, stratified by utilization cluster. This figure plots patient visit frequency for the two patients with highest and lowest average residuals in their utilization cluster for the year 2024. Each gray rectangle represents a patient, with visits of various types plotted in time across the year. Note that there are high density, but low frequency patients (left panel, rows 5-6), as well as patients who were hospitalized, but have low density data (right panel, rows 1-2). There is a clear association between high density and high visit frequency, but stratifying by utilization cluster allows us to see that that does not always hold true–utilization clusters and residuals are measuring different things.

**Figure 2.**
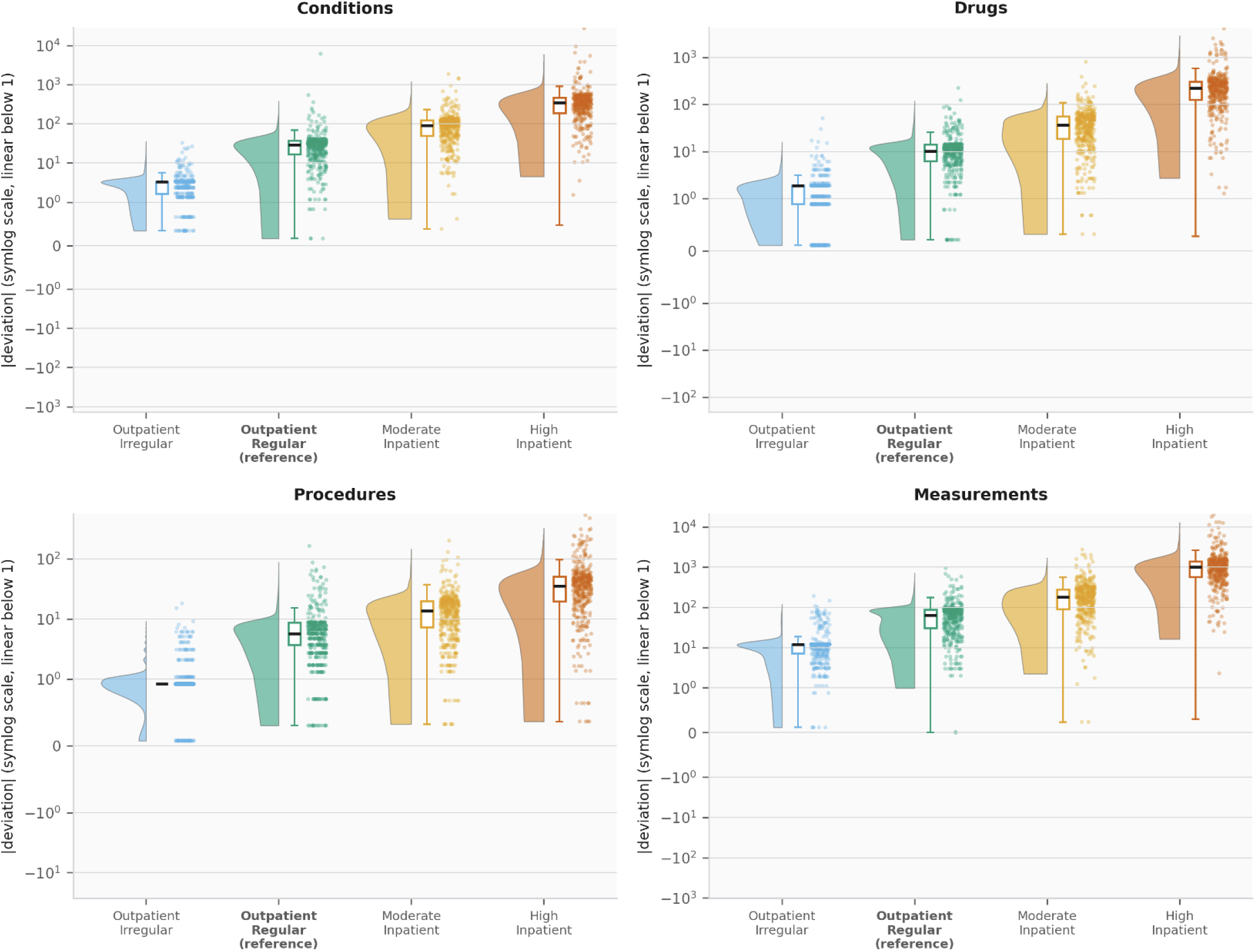
Within clusters, patient-year data density is heterogenous. The violin plots show how far patients stray from their cluster mean for each domain. Wider violins equate to more heterogeneity within that cluster. There is a clear relationship between cluster and degree of deviation from the cluster mean–e.g., the High Inpatient cluster shows the highest degree of deviation. However, the width of the violins illustrates that density can vary widely within a cluster, with even some High Inpatient cluster members having low density.

**Figure 3.**
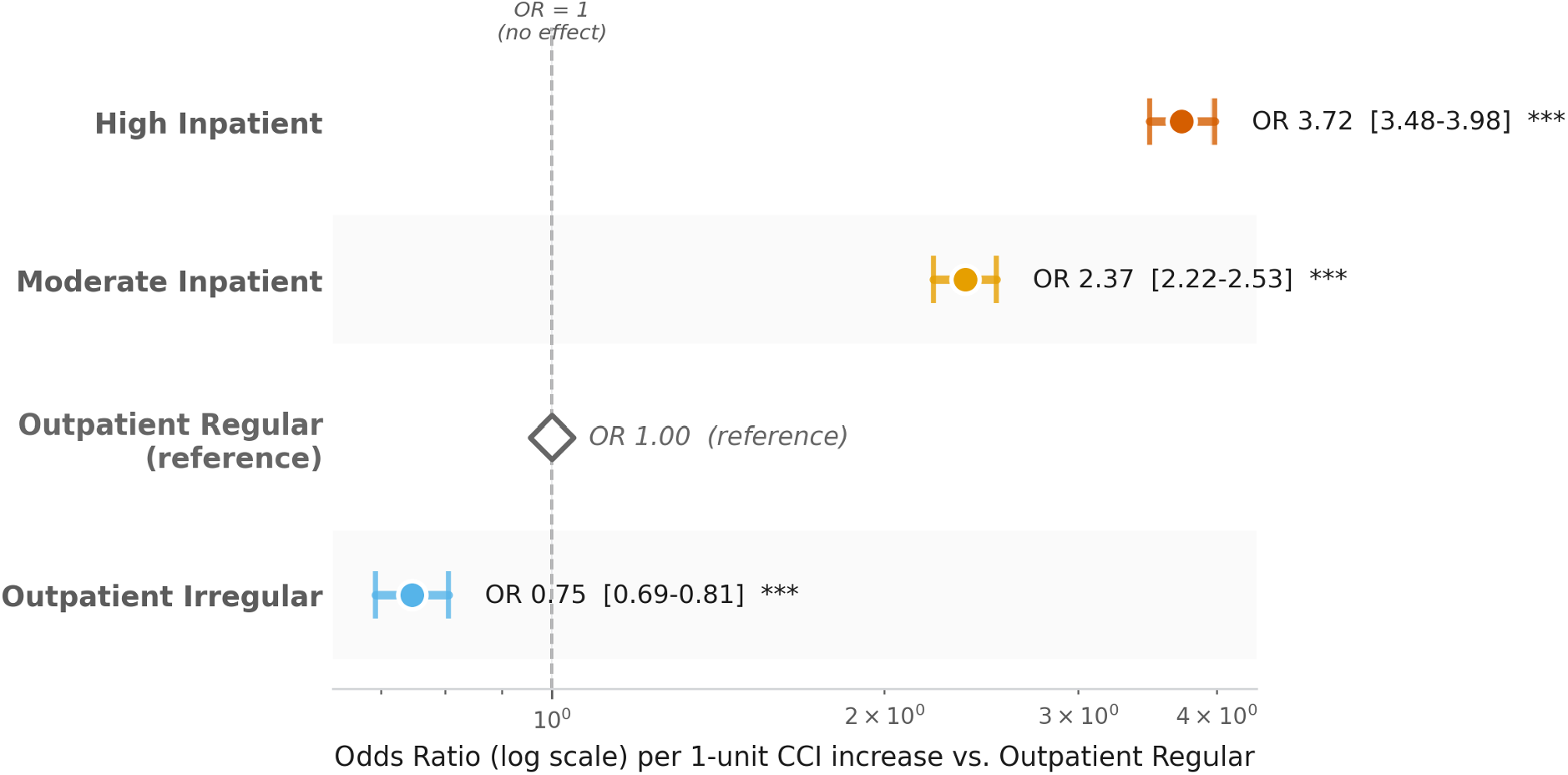
CCI is associated with cluster assignment. Each odds ratio reflects the multiplicative change in odds of belonging to that cluster vs Outpatient Regular for each 1-unit increase in cumulative CCI. Points to the right of OR = 1 (dashed line) indicate higher CCI predicts membership in that cluster vs Outpatient Regular. An OR gradient from low to high values confirms that CCI discriminates cluster membership in the expected clinical direction.

The EDI is intended as a covariate for use alongside other patient-level adjustments, not as an exposure or outcome. It is most useful in analyses where between-patient variation in EHR documentation density could confound or bias the estimate of interest, including matching, propensity score construction, regression adjustment, and sensitivity analyses. Adding the EDI to a model that already includes the CCI separates how sick a patient is, which the CCI captures, from how much we know about the patient, which the EDI captures.

Consider an analysis like our recent target trial emulation of Paxlovid for the prevention of hospitalization among adults with COVID-19.^9^ Here, we matched treated and untreated patients on baseline CCI to balance disease burden across arms. With the EDI available, the same analysis could additionally match or weight on baseline utilization cluster and mean cross-domain residual. This addresses a residual form of informative presence bias that CCI adjustment alone leaves uncorrected: in EHR data, exposure, outcome, and covariate ascertainment all scale with documentation density, and density is itself associated with treatment receipt.^1–3^ Including the EDI alongside the CCI would have addressed a residual-confounding limitation typical of EHR-based TTEs and reduced noise in the hospitalization estimate by absorbing the documentation-density component of outcome detectability.

Because the EDI is calculated on a per-year grain, the choice of which year (or years) to use depends on study design. For analyses with a single index date, we recommend using the EDI from the year immediately preceding the index date; this lag avoids contamination of the index year’s documentation by the event itself.

For analyses where exposure varies over time, the EDI can be carried as a time-varying covariate by joining the corresponding year’s EDI to each person-year of follow-up, mirroring how time-varying confounders are handled in Cox proportional hazards or generalized linear mixed models. When a single static covariate per patient is preferred, we recommend using the mean EDI across the study window, at the cost of losing within-patient variation across years.

Because documentation practices can vary widely across health care organizations, it is unlikely for the exact parameters of our pre-trained model to be fully generalizable across sites. It is our intention for the *method* to be generalizable, however, and as such we would strongly encourage investigators from another health system or multi-site collaborative wishing to use this work to first retrain the model on local data. Additionally, the EDI is built using structured EHR data only, and does not account for information sourced from clinical notes or other unstructured data sources. In future work, we hope to incorporate free text as an additional domain in our density vector.

The EHR is a rich source of real-world data, but its utility for research is inseparable from its characteristic unevenness. Patients differ not only in how sick they are, but in how much we know about them. The EDI addresses this gap by providing researchers with a per-patient, per-year characterization of documentation density that is distinct from disease burden as measured by the CCI or Elixhauser Index. By combining utilization-based clustering with within-cluster domain residuals, the EDI captures how much data a patient contributes to a given year of their record relative to patients with similar healthcare utilization patterns. Our analyses confirm that while disease burden and data density are associated, the relationship is weak enough that neither captures what the other measures. The EDI is not a replacement for established comorbidity measures, but a complement to them, intended for use as a covariate in analyses where documentation density may confound or bias the outcome of interest. We encourage researchers working with EHR data to consider incorporating the EDI alongside traditional comorbidity adjustment as a means of reducing a pervasive but often unaddressed source of bias in EHR data-driven studies.

## Data Availability

The randomly selected UNC Health records used to train the model described here are not publicly available due to HIPAA restrictions. However, we designed our model with the intention that it would be retrained on others data before use in another context. For this reason, we leveraged the OMOP common data model and have publicly posted our code (see Code Availability) to enable this work to be shared without requiring sensitive data sharing.The code used to train and deploy our model is available at https://github.com/abhatia08/data-density/, with a snapshot of the current state of the code available at https://doi.org/10.5281/zenodo.20706795.

https://github.com/abhatia08/data-density/

https://doi.org/10.5281/zenodo.20706795.

## Acknowledgements

The work described in this manuscript was supported by the National Center for Advancing Translational Sciences (NCATS), National Institutes of Health, through Grant Award Number UM1TR004406. The content is solely the responsibility of the authors and does not necessarily represent the official views of the NIH.

## Author Contributions

Study design: AB, SL, TM, EP; data curation and analysis: AB, SL, TM, EP; initial manuscript drafting: AB, TM, EP; manuscript review and editing: AB, SL, TM, EP; project leadership and funding acquisition: EP.

## Data Availability

The randomly selected UNC Health records used to train the model described here are not publicly available due to HIPAA restrictions. However, we designed our model with the intention that it would be retrained on others’ data before use in another context. For this reason, we leveraged the OMOP common data model and have publicly posted our code (see Code Availability) to enable this work to be shared without requiring sensitive data sharing.

## Code Availability

The code used to train and deploy our model is available at https://github.com/abhatia08/data-density/, with a snapshot of the current state of the code available at https://doi.org/10.5281/zenodo.20706795.

## References

1. Rusanov, A., Weiskopf, N. G., Wang, S. & Weng, C. Hidden in plain sight: bias towards sick patients when sampling patients with sufficient electronic health record data for research. BMC Med. Inform. Decis. Mak. 14, 51 (2014).

2. McGee, G., Haneuse, S., Coull, B. A., Weisskopf, M. G. & Rotem, R. S. On the nature of informative presence bias in analyses of electronic health records. Epidemiology 33, 105–113 (2022).

3. Weber, G. M. et al. Biases introduced by filtering electronic health records for patients with ‘complete data’. J. Am. Med. Inform. Assoc. 24, 1134–1141 (2017).

4. Charlson, M. E., Pompei, P., Ales, K. L. & MacKenzie, C. R. A new method of classifying prognostic comorbidity in longitudinal studies: development and validation. J. Chronic Dis. 40, 373–383 (1987).

5. Quan, H. et al. Coding algorithms for defining comorbidities in ICD-9-CM and ICD-10 administrative data. Med. Care 43, 1130–1139 (2005).

6. Elixhauser, A., Steiner, C., Harris, D. R. & Coffey, R. M. Comorbidity measures for use with administrative data. Med. Care 36, 8–27 (1998).

7. Drosdowsky, A. & Gough, K. The Charlson Comorbidity Index: problems with use in epidemiological research. J. Clin. Epidemiol. 148, 174–177 (2022).

8. Renson, A. & Bjurlin, M. A. The Charlson index is insufficient to control for comorbidities in a national trauma registry. J. Surg. Res. 236, 319–325 (2019).

9. Bhatia, A. et al. Effect of nirmatrelvir/ritonavir (Paxlovid) on hospitalization among adults with COVID-19: An electronic health record-based target trial emulation from N3C. PLoS Med. 22, e1004493 (2025).

